# A novel whole-blood stimulation assay to detect and quantify memory T-cells in COVID-19 patients

**DOI:** 10.1101/2021.03.11.21253202

**Authors:** William Mouton, Christelle Compagnon, Kahina Saker, Soizic Daniel, Xavier Lacoux, Bruno Pozzetto, Guy Oriol, Sophia Djebali, Franck Berthier, Jacqueline Marvel, Thierry Walzer, Karen Brengel-Pesce, Sophie Trouiller-Assant on behalf of COVID-ser study group

## Abstract

SARS-CoV-2 specific T-cells responses are essential for virus clearance. We present a novel and simple whole-blood assay allowing the detection of interferon-gamma-producing antiviral T-cells following peptide stimulation. We show that unlike neutralizing antibodies, antiviral memory T-cells persist at least 6 months in convalescent Covid-19 individuals.

## Introduction

As our understanding of the immune response against SARS-CoV-2 improves, it becomes clear that virus-specific T-cells are key players in the control of infection and that suboptimal T-cells response likely explains Covid-19 severity in many individuals [1]. SARS-CoV-2 specific T-cells responses often target epitopes conserved in different virus clades, including variants of concern [2], and even in other seasonal human coronaviruses (HCoV) [3]. Thus, memory T-cells may be more cross-protective than antibodies against different coronavirus infections. In addition, specific SARS-CoV-2 T-cells responses have been detected in convalescent donors without antibody responses [3] suggesting that, at least in some individuals, the measurement of T-cells responses may outperform serological measurements to uncover past infections. Thus, novel immunodiagnostics that measure cellular immune response to SARS-CoV-2 are needed to assess individual immune status and evaluate emerging vaccines in a robust and standardized manner, adapted to the clinical routine.

For this purpose, we designed a novel semi-automated whole-blood immune functional assay (WB IFA) to detect and quantify SARS-CoV-2-specific T-cells immunity. WB assays are rapid and simple assays that preserve all interactions between circulating immune cells, which likely reflects the *in vivo* situation [4]. Interferon-gamma (IFNg) secretion measurement following WB SARS-CoV-2 peptides stimulation was used to reveal SARS-CoV-2 specific T-cells, as previously described [5]. Using the IFA, we monitored T-cells immunity directed against different SARS-CoV-2 proteins including nucleocapsid (NC), membrane glycoprotein (MBGP) and receptor-binding domain (RBD) in a cohort of 129 Covid-19 convalescent healthcare workers (HCWs) previously infected 6 months before with SARS-CoV-2. We correlated these measurements with serological levels of antibodies (Abs) against SARS-CoV-2. We also assessed the specificity of the IFA assay for SARS-CoV-2 by performing measurements in 25 SARS-CoV-2 seronegative healthy volunteers (HV) and in 3 HCWs having experienced a previous documented infection by HCoV.

## Methods

### Study design

A prospective cohort study was conducted at the Hospices Civils de Lyon, France in HCWs with symptoms suggestive of SARS-CoV-2 infection. The clinical study was registered on ClinicalTrial.gov (NCT04341142) [6]. Written informed consent was obtained from participants and the study was approved by the national review board for biomedical research (Comité de Protection des Personnes Sud Méditerranée, ID-RCB-2020-A00932-37). Nasopharyngeal swabs collected at inclusion were tested with the BIOFIRE^®^ Respiratory Panel (bioMérieux,). At 6 months post-infection, blood sampling was performed on 129 HCWs who were SARS-CoV-2 positive and 3 HCWs who were HKU-1 or NL-63 positive. According to French procedures, a written non-opposition to the use of donated blood for research purposes was obtained from HV. The donors’ personal data were anonymized before transfer to our research laboratory. We obtained approval from the local ethical committee and the French ministry of research (DC-2008-64) for handling and conservation of these samples.

### Serological investigations

The presence of anti-SARS CoV-2 Ab was evaluated on serum samples using the Wantai Ab assay that measures total Abs against the RBD of the S protein and the bioMérieux Vidas^®^ assay that measures IgG to the RBD. Positivity was established according to the threshold value recommended by each manufacturer. The neutralizing antibody (NAb) titres were determined by a virus neutralization test (VNT) using live SARS-CoV-2 virus as previously described [7].

### T-cells response after WB stimulation against SARS-CoV-2 peptides

Fresh blood collected in heparanized tubes was stimulated for 22 hours at 37°C under 5% of CO_2_ with peptide pools targeting RBD (46 peptides), MBGP (53 peptides) or NC (102 peptides) (bioMérieux, France) diluted in IFA solution (bioMérieux, France). The IFA solution was used as a negative control and a mitogen as a positive control. The peptides (15-mer) encompassed the whole protein sequence and overlapped by 5-residues. The concentration of IFNg in the supernatant was measured using the VIDAS automated platform (VIDAS^®^ IFNg RUO, bioMérieux). The measuring range was 0.08 −8 IU/mL and IFA positivity thresholds were defined at 0.08 IU/mL. The IFNg response was defined as positive when the IFNg concentration of the test was above threshold and the negative control was below threshold or when the IFNg concentration of the test minus IFNg concentration of the negative control was above threshold. All positive controls were ≥8 IU/mL.

### Statistical analysis

For T-cells responses, comparisons were performed using non-parametric Kruskal-Wallis test and Dunn’s multiple comparisons test; the overall, positive and negative percent agreements were determined between T-cells response and serological assays or VNT as previously described [8]. Statistical analyses were conducted using GraphPad Prism^®^ software (GraphPad software, La Jolla, USA) and R software. A p-value ≤ 0.05 was considered as statistically significant.

## Results

We first monitored SARS-CoV-2 specific Abs using the Wantai total Ab kit in the three different groups of participants (HV, or convalescent HCWs with previous infection by SARS-CoV-2 or HCoV occurring 6 months before). These Abs were undetectable in the sera from the 3 HCoV positive HCWs and from the 25 HV, confirming the absence of SARS-CoV-2 previous infection. By contrast, they were detected in all 129 HCWs with documented SARS-CoV-2 infection 6 months before. Among them, we observed that 82.9% (n=107) were positive for IgG anti-SARS-CoV-2 RBD detection (bioMérieux VIDAS) and that only 48.8% (n=63) had neutralizing Abs (Nabs, Supplementary Table 1).

We then monitored the T-cells response against MBGP, RBD and NC peptides using the IFA WB assay for the different patients’ groups. A positive IFNg release was detected for 89.1% (n=115/129; median 0.51 [0.24-1.49] IU/mL), 62.4% (n=58/93; median 0.12 [0.17-0.34] IU/mL) and 97.9% (n=92/94; median 1.20 [0.60-2.48] IU/mL) of SARS-CoV-2 positive HCWs after stimulation with MBGP (Figure 1A), RBD (Figure 1B) and NC (Figure 1C) SARS-CoV-2 peptides respectively. No or very low IFNg release (<0.12 IU/mL) was detected for HV and HCoV upon stimulation with MBGP or RBD (Figure 1A et 1B). Interestingly, two of the three HCoV positive HCWs (0.45 [0.10-0.79] IU/mL), as well as 44% (n=11/25, 0.20 [0.13-0.32] IU/mL) of the HV showed a positive IFNg response upon stimulation with NC (Figure 1C), confirming that HCoV-specific T-cells cross-reacted with some of the SARS-CoV-2 NC peptides that were used [9].

**Figure 1:**
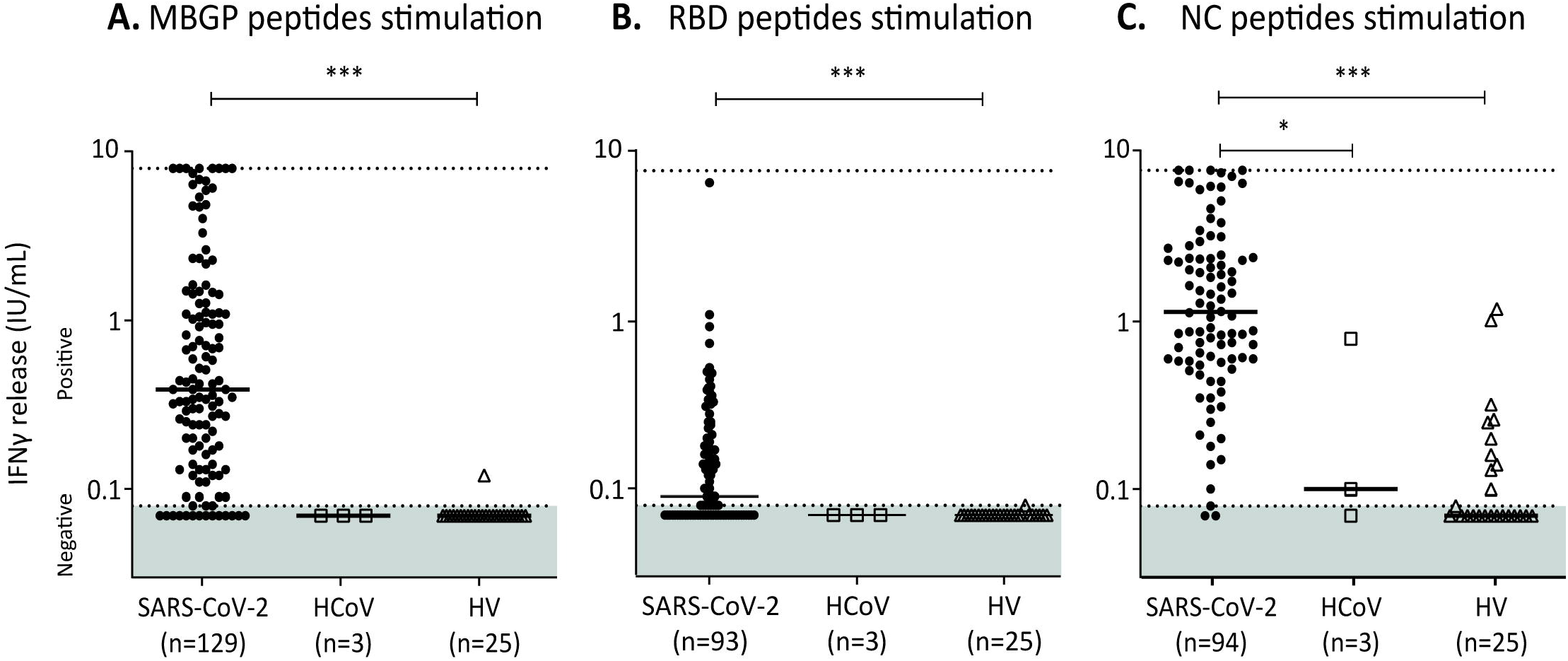
IFNγ release after in vitro WB stimulation using SARS-CoV-2 peptide pools. IFN1 levels in WB samples from 129 SARS-CoV-2 convalescent HCWs (dot), 3 HCoV convalescent HCWs (square) and 25 controls (HV) (triangle). IFN1 secretion was measured after a 22-hour stimulation using **(A)** MBGP peptides, **(B)** RBD peptides and **(C)** NC peptides. Statistical differences were inferred using non-parametric Kruskal-Wallis test and Dunn’s multiple comparisons test. <u>Abbreviations:</u> HCoV: seasonal human coronaviruses; HCWs: healthcare workers; HV: healthy volunteers; IU: international unit; MBGP: membrane glycoprotein; NC: nucleocapsid; RBD: receptor binding domain.

The overall percent agreement between T-cells responses against MBGP or RBD IFA and positive IgG anti-SARS-CoV-2 RBD (BioMérieux IgG assay) was of 78.29% and 70.97% respectively. Among the 22 Covid-19 convalescent HCWs with undetectable anti-SARS-CoV-2-RBD IgG, only 4 did not have MBGP-specific T-cells, which resulted in a low negative percent agreement between cellular and humoral responses (18.18%). Moreover, among the 66 convalescent HCWs who lacked neutralizing Abs at 6 months post-infection, 81.8% (n=54) had MBGP-specific T-cells (detailed in supplementary Table 1). These results suggest that, in the case of undetectable nAbs against SARS-CoV-2, a T-cells response can still be observed.

## Concluding remarks

Here, we presented a novel WB IFA assay that is highly efficient in detecting SARS-CoV-2 specific T-cells responses, especially those directed against MBGP. IFNg-release quantification following WB stimulation could thus be proposed as a suitable and rapid option for assessing the presence of a long-term specific cellular response after natural SARS-CoV-2 infection.

In our cohort of Covid-19 convalescent HCWs, we observed that about 90% of patients had a detectable T-cells response after SARS-CoV-2 peptide stimulation at 6 months post symptoms, in accordance with recent results [10]. Thus, our results suggest that WB IFA stimulation assays can be used to detect memory T-cells in convalescent patients, even at late phases post infection, extending previous findings by *Murugesan* et al. [5] who used a similar approach in recently-infected patients by SARS-CoV-2. Moreover, we confirmed that IFNγ secretion was a robust read-out to assess SARS-CoV-2 memory T-cells responses over time [11]. Yet, peptides derived from distinct viral proteins showed a different capacity to trigger a T-cells response in patients. The highest positivity was observed after stimulation with NC peptides. These peptides could even induce IFNg secretion by HCoV-convalescent HCWs and HV T-cells, suggesting cross-reactivity between seasonal HCoV and SARS-CoV-2 and potential pre-existing immunity in HV, as previously described [9,12,13]. This also suggests that stimulation using NC peptides encompassing the whole NC protein should not be used as a specific diagnostic tool of SARS-CoV-2 immunity. By contrast, MBGP peptides seem to be more appropriate to reveal SARS-CoV-2 specific memory T-cells. We also confirmed previous findings regarding the relative short half-life of NAbs and anti-SARS-CoV-2 IgGs [7]. Thus the measurement of T-cells responses may outperform serological measurements to uncover past infections. We acknowledge that our study has some limitations: (i) the WB IFA assay was not able to differentiate between CD8+ and CD4+ T-cells responses, (ii) it does not determine whether memory T-cells are protective against SARS-CoV-2 re-infection and (iii) it was not yet evaluated in vaccinated subjects.

## Supporting information

Supplementary Table 1 and results

## Data Availability

Data not available due to commercial restrictions

## Acknowledgments

We thank all the personnel of the occupational health and medicine department of Hospices Civils de Lyon who contributed to the samples collection. We thank Karima Brahami and all members of the clinical research and innovation department for their reactivity (DRCI, Hospices Civils de Lyon). A special thanks to Virginie Pitiot and PMO team for their help in patient enrolment and clinical data collection.

## No conflict of interest

KS, BP, SD, JM, TW and STA

