## Supplementary Table 1 and results for "A novel whole-blood stimulation assay to detect and quantify memory T-cells in COVID-19 patients"

**Supplementary Table 1 –** Association of SARS-CoV-2 humoral and cellular response

| **Manufacturer** | **bioMérieux** | **Virus neutralization test** |
| --- | --- | --- |
| **(platform)** | **(Vidas®)** | **(VNT)** |
| *Assay name* | SARS-CoV-2 IgG | PRNT_50_ <20 |
| *Assay type* | ELFA | / |
| *Antigen* | RBD | / |
| *Positive, n* ***(%)*** | 107 (**82.9**) | 63 (**48.8**) |
| *Negative, n* ***(%)*** | 22 (**17.1**) | 66 (**51.2**) |
| **Overall, Negative and Positive Percent Agreement with IFA MBGP ; T-cell responders 115/129 (89.1%)** | | |
| *OPA [95%CI]* | 78.3 [70.4-84.5] | 56.6 [48.0-64.8] |
| *PPA [95%CI]* | 90.7 [83.7-94.8] | 96.8 [89.1-99.1] |
| *NPA [95%CI]* | 18.2 [7.3-38.5] | 18.2 [10.7-29.1] |
| **Overall, Negative and Positive Percent Agreement with IFA RBD ; T-cell responders 58/93 (62.4%)** | | |
| *OPA [95%CI]* | 71.0 [61.1-79.2] | 62.4 [52.2-71.5] |
| *PPA [95%CI]* | 69.6 [58.8-78.7] | 72.6 [59.1-82.9] |
| *NPA [95%CI]* | 78.6 [52.4-92.4] | 50.0 [35.5-64.5] |
| **Overall, Negative and Positive Percent Agreement with IFA NC ; T-cell responders 92/94 (97.9%)** | | |
| *OPA [95%CI]* | 81.9 [72.9-88.4] | 51.1 [41.1-60.9] |
| *PPA [95%CI]* | 97.5 [91.2-99.3] | 100 [92.3-100] |
| *NPA [95%CI]* | 0 [0-20.4] | 4.2 [1.2-14.0] |

Abbreviations: Ab: antibody; CI : confidence interval; HCWs : healthcare workers; IFA: immune functional assay; IgG : Immunoglobulin G; MBGP: membrane glycoproteins; N/A: not applicable; NC: nucleocapsid; NPA: negative percent agreement; OPA: overall percent agreement; PPA: positive percent agreement; PRNT : plaque reduction neutralization test; RBD: receptor binding domain.

**Supplementary Results**

1. **Demographic and clinical characteristics of HV and coronavirus positive HCWs**

We monitored anti-SARS-CoV2 T-cells response using IFA in HV (n=25) and positive HCWs with SARS-CoV2 (n=129) or HCoV (n=3) infection at six months post-symptoms [7]. Of note, none of HCoV positive HCWs and HV were seropositive for SARS-CoV-2 at inclusion. Among the patients, 20/129 (16%) SARS-CoV-2 positive HCWs were male (sex ratio, 0.18) with a median age of 41 years (21-62 years) and 1/3 (33%) HKU-1 (n=1)/NL-63 (n=2) positive HCWs were male (sex ratio, 0.5) with a median age of 22 years (21-23 years), while 13/25 (52%) healthy volunteers were male (sex ratio, 1.08) with a median age of 42 years (19-68 years).
